# Mapping the Existing Body of Knowledge on New and Repurposed TB Vaccine Implementation: A Scoping Review

**DOI:** 10.1101/2024.01.18.24301490

**Authors:** Joeri S. Buis, Degu Jerene, Agnes Gebhard, Roel Bakker, Arman Majidulla, Andrew D. Kerkhoff, Rupali Limaye, Puck T. Pelzer

## Abstract

There is global consensus on the urgent need for a safe and effective TB vaccine for adults and adolescents to improve global TB control, and encouragingly, several promising candidates have advanced to late-stage trials. Significant gaps remain in understanding the critical factors that will facilitate the successful implementation of new and repurposed TB vaccines in low-and middle-income countries (LMICs), once available. By synthesizing the existing body of knowledge, this review offers comprehensive insights into the current state of research on implementation of these adults and adolescent vaccines. This review explores four key dimensions: (1) epidemiological impact, (2) costing, cost-effectiveness, and/or economic impact, (3) acceptability, and the (4) feasibility of implementation; this includes implementation strategies of target populations, and health system capabilities. Results indicate that current research primarily consists of epidemiological and costing/cost-effectiveness/economic studies in India, China, and South Africa, mainly modelling with M72/AS01, BCG revaccination, and generic vaccines. Varying endpoints, vaccine efficacies, and vaccination coverages were used. Globally, new, and repurposed TB vaccines are estimated to save millions of lives. Economically, these vaccines also demonstrate promise with expected cost-effectiveness in most countries. Projected outcomes were dependent on vaccine characteristics, target population, implementation strategy, timing of roll out, TB burden/country context, and vaccination coverage. Potential barriers for vaccine acceptability included TB-related stigma, need for a second dose, and cost, while low pricing, community and civil society engagement and heightened public TB awareness were potential enablers in China, India, and South Africa. Potential implementation strategies considered spanned from mass campaigns to integration within existing vaccine programs and the primary target group studied was the general population, and adults and adolescents. In conclusion, future research must have broader geographical representations to better understand what is needed to inform tailored vaccine programs to accommodate diverse country contexts and population groups to achieve optimal implementation and impact. Furthermore, this review underscores the scarcity of research on acceptability of new and repurposed TB vaccines and their delivery among potential beneficiaries, the most promising implementation strategies, and the health system capabilities necessary for implementation. The absence of this knowledge in these areas emphasizes the crucial need for future research to ensure effective TB vaccine implementation in high burden settings worldwide.

## Introduction

Tuberculosis (TB) remains a persistent global health challenge, imposing a significant burden on communities worldwide, despite substantial health efforts, including the availability of effective treatment, and widespread use of the Bacillus Calmette–Guerin (BCG) vaccine. In 2022 alone, 10.6 million people fell ill with TB, with approximately 1.3 million deaths^1^. Of all reported TB patients, 99% were in low-and middle-income countries (LMICs^1^). India, Indonesia, China, the Philippines, Pakistan, Nigeria, Bangladesh, and the Democratic Republic of Congo, account for approximately two-thirds of all patients worldwide. The development of new and repurposed tuberculosis (TB) vaccines is a promising approach with the potential to transform TB prevention, aiding (LMICs) in achieving the World Health Organization (WHO)’s End TB targets^1^. The WHO has identified new and repurposed vaccines as a cornerstone of the WHO End TB Strategy, with the ultimate goal to build a world without TB^1^. Effective implementation will be pivotal for maximizing public health impact once new and/or repurposed vaccines are available.

Currently, the BCG vaccine is the only licensed vaccine against TB. First employed in 1921, the vaccine is one of the most widely used vaccines globally, with 80% of all countries recommending BCG vaccination for neonates^2^, and a coverage of >80% on average. BCG vaccination primarily prevents severe forms of TB in young children and multiple limitations have been identified: varying effectiveness, contraindication in people who are living with HIV/AIDS (PLHIV), and the lack of protection against reactivation of TB. Additionally, there is little evidence that it prevents the development of TB in adults and adolescents^3^. These are important limitations as adults and adolescents account for about 90% of TB incidence and TB is the leading cause of death from a single infectious agent in PLHIV^1,4^.

To fill the critical research and development gap in TB vaccines, there has been an increase in clinical trials evaluating the effectiveness and safety of TB vaccine candidates that target adults and adolescents. Approximately 17 TB vaccine candidates are in the clinical development pipeline, of which six have progressed to phase 3 development and are intended for use in adults and adolescents^4,5^. These include MIP, VMP1002, GamTBvac, MTBVAC, BCG (re)vaccination, and M72/AS01E. Of these, promising phase 2b results were published for the M72/AS01E vaccine, revealing a vaccine efficacy of 49.7% (90% confidence interval [CI]=12.1 to 71.2 and 95% CI=2.1 to 74.2) against development of TB disease over a three year follow-up period ^5^. For BCG revaccination in adolescents between the ages of 12-17 years, vaccine efficacy in preventing TB infection was estimated at 45.4% (95% CI, 6.4 to 68.1)^6^. While clinical trials are instrumental in evaluating the safety and efficacy of TB vaccine candidates, evidence on vaccine implementation beyond the clinical trial setting is critical to achieving public health impact. There is an urgent need to understand how to effectively implement and scale-up vaccination programs to achieve optimal public health impact, following the approval of new and repurposed TB vaccines.

In the last few years, evidence has accumulated regarding the potential impact of new and repurposed TB vaccines. Modelling studies have shown that a new TB vaccine with an efficacy of at least 50% for preventing the development of TB disease, a faster and broader impact could be achieved targeting adults and adolescents rather than infants and children^7^. However, Harris et al. argue that some approaches used in the modelling – such as campaigns specifically targeting uninfected people – may not be feasible due to the high cost and inaccuracies of available tuberculin skin tests to exclude TB infection^7^.

Two recent documents, the Global Roadmap for Research and Development for TB Vaccines and the Evidence Considerations for Vaccine Policy (ECVP), identify key actions and considerations to prepare for the effective implementation of a new and repurposed TB vaccine, once approved^8,9^. Crucial TB vaccine implementation components to realizing successful TB vaccination programs for adults and adolescents in LMICs include understanding the TB burden, the public health and economic impact of TB vaccination in target populations, developing and implementing strategic plans for TB vaccination, strengthening healthcare capacity and infrastructure, enhancing public awareness, and increasing acceptance among the public^8,9^.

A comprehensive review and synthesis of the existing literature on best practices regarding acceptability and feasibility of implementing new or repurposed TB vaccines for adults and adolescents has not yet been undertaken, nor has an overview of projected epidemiological impact and costing, cost-effectiveness and economic impact.^2^ This work would offer critical insights to understanding what will be needed to prepare for and contribute to the effective launch and scalable delivery of a licensed TB vaccine upon availability, including the identification of knowledge gaps.

Therefore, the objective of this scoping review is to assess and synthesize existing research and evidence on TB vaccine implementation in adults and adolescents in LMICs, regarding projected epidemiological impact, economic impact, acceptability, and feasibility, and identify priority areas for future research to prepare for deployment of new and repurposed TB vaccines. The specific questions included:

What is the existing body of knowledge on the:

1. epidemiological impact of new and repurposed TB vaccines targeting adults and/or adolescents as estimated by modelling studies?
2. costing, cost-effectiveness, and/or economic impact of new and repurposed TB vaccines targeting adults and/or adolescents as estimated by modelling studies?
3. acceptability of new and repurposed TB vaccines targeting adults and/or adolescents?
4. implementation feasibility of vaccination programs using new and repurposed TB vaccines, targeting adults and/or adolescents?

a. the potential implementation strategies for new and repurposed TB vaccines?
b. the existing body of knowledge on health system readiness for the implementation and scale up of a new and repurposed TB vaccine?

## Methods

### Study design overview

We employed a narrative synthesis approach^10^, sourcing literature from PubMed, Medrxiv, PLOS journals, and expert suggestions. Articles were selected using a systematic method, and data extraction and analysis were carried out iteratively using the Roadmap^8^ and ECVP^9^.

### Search Methods

The narrative synthesis approach involved three systematic search components: (1) database search, (2) search on internet sites, and (3) identification of relevant articles by the team. The search strings for the database queries combined the following concepts: TB, TB vaccination, and vaccine preparedness. A combination of Medical Subject Headings (MeSH) terms and separate keywords relating to the research questions were included to create a broad and inclusive search strategy. All concepts included variations of the keywords. Filtering removed non-human, infant/children, clinical, and immunological studies, limiting to English articles published between 2013-2023. Using the search strings, article retrieval was carried out on 30^th^ April 2023. The detailed search strings are provided in Annex A.

1. Databases: Searches were conducted using PubMed and Medrxiv.
2. Internet sites: PLOS journals were identified as having relevant literature related to this topic, however not all PLOS journals were indexed in PubMed or Medrxiv at the time of conducting the search. Therefore, a quick literature search was conducted within the PLOS journal.
3. Identification of additional literature: to accommodate the identification of articles currently not covered within the above search components, additional literature identified by the internal team of experts on TB vaccine readiness were also included.

### Inclusion and exclusion criteria

Scientific literature (including preprints) related to the implementation of a new and repurposed TB vaccines for adults and adolescent was included (Table 1). We focused on studies conducted in or concentrating on LMICs and we focused on articles published between 2013 and 2023. Expanding the timeframe was not anticipated to yield additional relevant articles, given that the development of TB vaccine candidates has been a more recent occurrence. Studies focusing on vaccine candidates moving to/are in phase 3 trials were included as were generic vaccines that align with the WHO Preferred Product Characteristics (PPC)^8^. Literature covering other types of vaccines, (clinical/non-human) trials, and immunological studies were excluded.

**Table 1.** Inclusion and exclusion criteria for selection of literature.

| Inclusion criteria | Exclusion criteria |
| --- | --- |
| Type of study: Studies (including preprints) on the preparedness and implementation of new and repurposed TB vaccines | Non-human trials and immunology studies |
| Country category: Low- and middle-income countries | TB vaccines trials on efficacy/ effectiveness, and safety |
| Age group: Adults and/or adolescents, with the specific age as defined in the study | Children, infants, neonates |
| Language: English only | BCG for neonates |
| Conducted between 2013-2023 | Books, guidelines, frameworks, roadmaps, presentations |

Studies on BCG vaccines (other than explicitly related to a revaccination strategy) were also excluded as were studies focused on the efficacy/effectiveness, and safety of new and repurposed TB vaccines. The primary focus of our review was on collecting primary data regarding the implementation of new and repurposed TB vaccines. Consequently, we chose not to include a grey literature search, as our expectation was that this type of literature might not predominantly contain primary data.

### Selection of articles

The literature assessment occurred in a sequential process. Initially, articles from PubMed and Medrxiv were imported into Zotero, where their abstracts and titles were evaluated for relevance. Simultaneously, PLOS journal articles were examined directly on its website, and those meeting the relevancy criteria were individually added to Zotero, unless already detected through the other searches. Subsequently, a comprehensive full-text screening was conducted to in-or exclude articles, while removing duplicates. In instances where articles reported secondary results, the initial paper was added. JSB undertook the initial selection, screening, and assessment. PTP checked a random sample of articles to ensure consistency in the inclusion process. After title and abstract screening, JSB checked articles for their relevance through a full-text assessment. JSB consulted PTP to discuss articles for potential inclusion in cases of uncertainty. After full-text inclusion, the data were subtracted and collected in an extraction sheet using Excel 2023. A thorough assessment (snowballing) of the references was performed on both the articles cited within the identified reviews and the final selected articles to ascertain any potentially relevant literature that was not captured within the initial search strategies. Similarly, any articles identified through the review of references and reviews followed the same structured inclusion process, as did any additional articles identified by the team.

### Data extraction and analysis

The AIGHD/EDCTP Roadmap and WHO’s Evidence Considerations for Vaccine Policy^8,9^ provided key implementation factors for ensuring public health impact in shaping our analytical framework. Four pivotal implementation research components were evaluated: (1) Epidemiological impact (e.g. cases and deaths averted by vaccine), (2) Economic impact – on health systems, societies, macroeconomic-, value for money (e.g. cost per death or DALY averted, budget impact and impact on equity and social protection, including costing and cost-effectiveness, (3) Acceptability by potential target population (i.e. the likely acceptability, including barriers and enablers, of the vaccine in the target populations and other key stakeholder groups), and (4) Implementation feasibility by potential target population (i.e. practicality of vaccine implementation, considering the logistics, delivery, and program related considerations). We adapted the conceptual framework from Atun ^11^ based on these key components (Figure 1).

**Figure 1.**
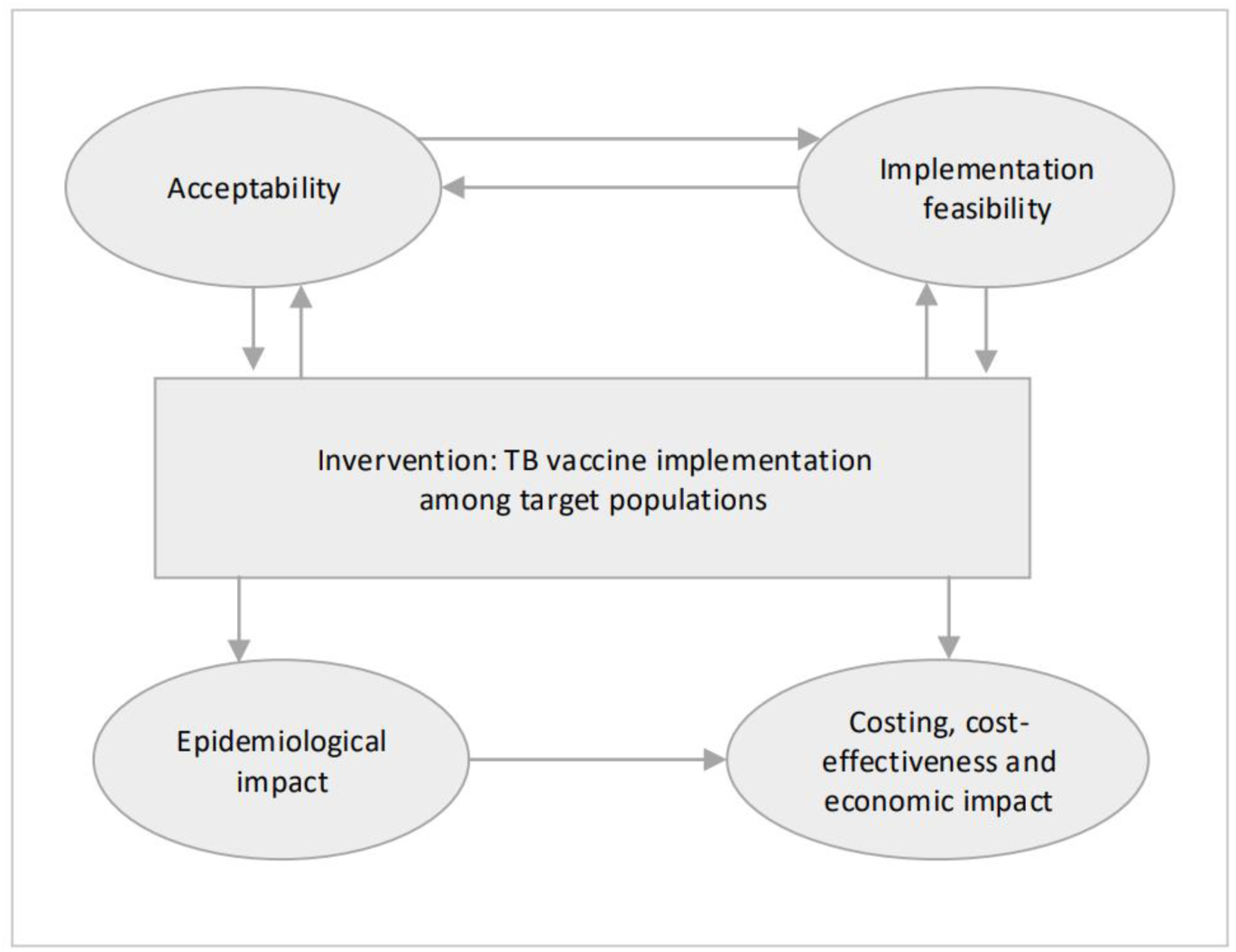
Conceptual framework of factors influencing implementation of TB vaccination among adults and adolescents^9,11^.

We listed the number of studies conducted within each component in an Excel sheet by country and evaluated each study based on the four components. Results were reported for countries separately, outlining the vaccine characteristics, anticipated vaccination coverage, and time spans of the projected TB vaccine roll out. Implementation feasibility was further assessed through the lens of WHO’s health systems building blocks^12^ : health service delivery, health workforce, health information systems, accessibility, health financing, and leadership and governance.

## Results

### Articles output

We identified 1,807 articles of which 682 were from PubMed and 1,125 from Medrxiv (Figure 2). Following title and abstract screening of these initial articles and after removal of duplicates, 81 articles were included for full-text screening. The search strategy for PLOS journals resulted in the identification of two additional articles, which were included for full-text screening. Of the 83 articles for full-text inclusion that were reviewed, 66 articles did not meet the inclusion criteria of which seven articles referenced other articles with primary data. Five articles were identified through snowballing and one additional article was added by the team, resulting in 23 total articles for inclusion. Of the 23 articles, 18 were peer-reviewed, and 5 were scientific preprints.

**Figure 2.**
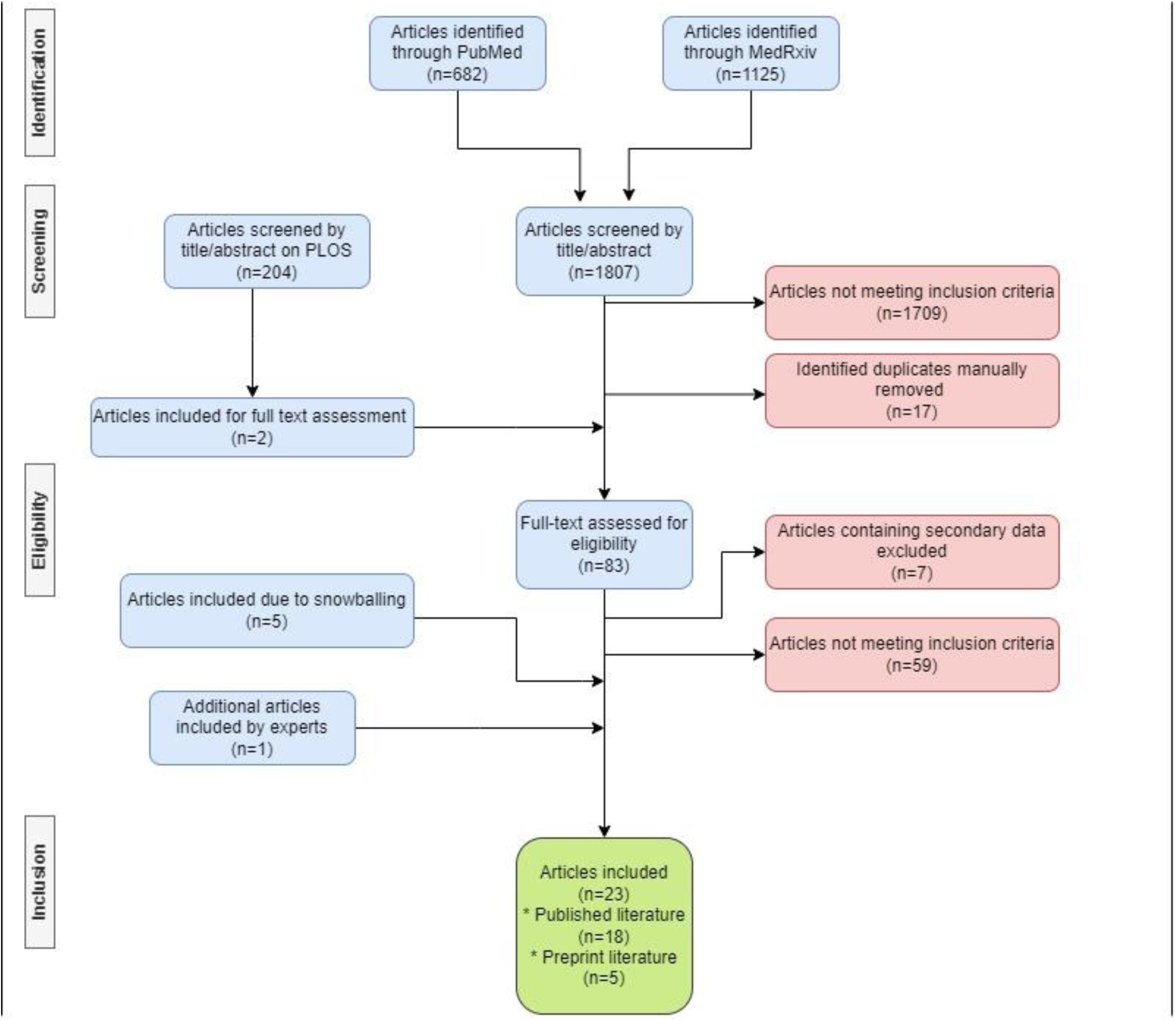
Flow diagram of article selection.

### Study characteristics

A total of 23 articles were included in the review. Table 2 provides an overview of the number of included studies per country, vaccine readiness focus, vaccine candidate or profile, and vaccine endpoints. Most of the research was conducted in/ or focused on India (9 studies), followed by high burden LMICs in general (7 studies), and articles specific by country: South Africa (7 studies), China (6 studies), Indonesia (1 study), and Cambodia (1 study). Among the key research topics on TB vaccine implementation, epidemiological impact was the most frequently assessed topic (16 studies), followed by costing, cost-effectiveness, and/or economic impact (11 studies). One article addressed acceptability within the context of implementation feasibility, with a specific focus on exploring implementation strategies and health system readiness while five articles assessed implementation strategies. Various vaccine candidates and profiles were covered in the literature, including M72 /AS01e (8 studies), BCG revaccination (5 studies), and generic vaccines (15 studies). Endpoints for the generic vaccines, most frequently encompassed prevention of disease (PoD) (14 studies), while prevention of infection (PoI) (6 studies) was less common; no study modelled a prevention of recurrence (PoR) endpoint (see table 3).

**Table 2.**
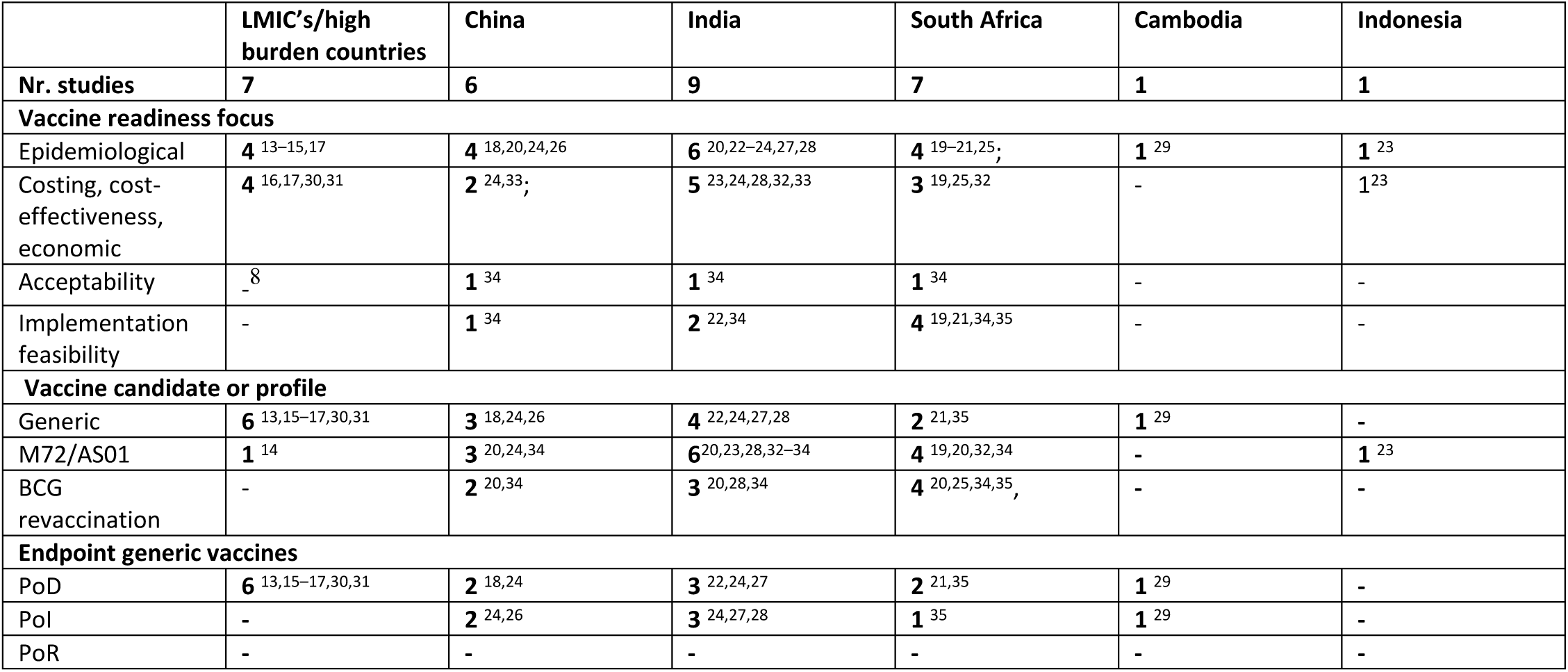
number of included studies per country, vaccine readiness focus, vaccine candidate and profile, and vaccine endpoint.

**Table 3.** Overview characteristics included articles.

| First author, year, | Type of literature | Type of study | country | Type of vaccine | Target population | Implementation strategy | Health impact | Economic impact | Acceptability | Feasibility |
| --- | --- | --- | --- | --- | --- | --- | --- | --- | --- | --- |
| Pelzer, 2021 | white | Qualitative | China, India, South Africa | BCG revax & MS72 /AS01E | various; adolescents, general population, elderly, | mass campaigns, school program, routine immunization program, |  |  | x | x |

<sup>8</sup> No studies identified.

|  |  |  |  |  |  |  |  |  |  |  |
| --- | --- | --- | --- | --- | --- | --- | --- | --- | --- | --- |
|  |  |  |  |  | healthcare workers, high risk contacts, socially vulnerable groups, biological high-risk groups, | workplace, health visitations |  |  |  |  |
| Harris, 2019 | white | Modelling | China | Generic vaccine (POD) | elderly & adolescents | yearly routine vaccination; catch up campaigns for specific age groups | x |  |  |  |
| Hatherill, 2016 | white | Supplement | South Africa | Generic vaccine (POD/POI) and BCG revax | Healthcare workers | not specific |  |  |  | x |
| Harris, 2022 | white | Modelling | India, South Africa | MS72/AS01E | adolescents | routine vaccination |  | x |  |  |
| Shrestha, 2017 | white | Modelling | South Africa | Generic vaccine (POD) | miners and mine-labor-sending community | not specific | x |  |  | x |
| Dye, 2013 | white | Modelling | South Africa | BCG revax | adolescents (teenagers) | not specific | x | x |  |  |
| Arinaminpathy, 2022 | Pre-print | Modelling | Countries USAID TB portfolio | Generic vaccine (POD) | general population | not specific | x |  |  |  |
| Liu, 2017 | white | Modelling | China | Generic vaccine (POI) | general population | mixed vaccination: constant vaccination among new and repurposed pulse vaccination strategy among all other age groups through government agencies and companies | x |  |  |  |
| Fu, 2021 | white | Modelling | high RR-TB incidence countries | MS72/AS01E | adults, adolescents | routine vaccination and catch-up campaigns amongst those over 15-years old | x |  |  |  |
| Clark, 2023a | preprint | modelling | India | BCG revax, MS72 /AS01E, generic vaccine (POI) | general adults/adolescents | combination of routine vaccination, mass campaigns and a repeat campaign | x | x |  |  |
| Portnoy, 2023 | white | modelling | 105 LMIC | Generic vaccine (POD) | general adults/adolescents | routine vaccination and a onetime vaccination campaign |  | x |  |  |
| Weerasuriya, 2021a | white | modelling | China, India | Generic vaccine (POD & POI) | early adolescents | annual routine vaccination, co-delivered with HPV vaccine; mass campaigns | x | x |  |  |
| Clark, 2023b | white | modelling | 105LMICs | Generic vaccine (POD) | adults/adolescents | routine vaccination; mass campaign | x |  |  |  |
| Portnoy, 2022a | preprint | modelling | 105LMICs | Generic vaccine (POD) | general adults/adolescents | an initial mass vaccination campaign followed by routine vaccinations |  | x |  |  |
| Portnoy, 2022b | preprint | modelling | 105LMICs | Generic vaccine (POD) | general adults/adolescents | Routine vaccination plus a one-time vaccination campaign |  | x |  |  |
| Harris, 2020 | white | modelling | China, India, South Africa | BCG revax & MS72 /AS01E | general adults and adolescents | routine vaccination and mass campaigns | x |  |  |  |
| Weerasuriya, 2021b | white | modelling | China, India | MS72/AS01E | early adolescents | mass vaccination |  | x |  |  |
| Awad, 2020 | white | modelling | India | Generic vaccine (POD & POI) | people with diabetes | not specific | x |  |  |  |
| Renardy, 2019 | white | modelling | Cambodia | Generic vaccine (POD & POI) | general adults/adolescents | not specific | x |  |  |  |
| Knight, 2014 | white | modelling | LMIC's | Generic vaccine (POD) | general adults/adolescents | school and mass vaccination programs | x | x |  |  |
| Shrestha, 2016 | white | modelling | India | Generic vaccine (POD) | general adults/adolescents | yearly routine vaccination and 10y-periodic revaccination | x |  |  | x |
| Silva, 2021 | preprint | modelling | India, Indonesia | MS72/AS01E | general adults | not specific | x | x |  |  |
| Jayawardana, 2022 | white | qualitative +modelling study | South Africa | MS72/AS01E | adults/adolescents + PLHIV adults | adults/adolescents : 1. mass + routine and 2. 2x mass. PLHIV adults: 1. mass & routine, 2. 2xmassadults | x | x |  | x |

### Epidemiological and Costing, Cost-effectiveness, and/or Economic Modelling Studies

A total of 21 studies estimated the epidemiological and/or costing, cost-effectiveness, and/or economic benefits of new and repurposed TB vaccines as shown in table 4 & 5. The studies were conducted across multiple LMICs, with India (8 studies) leading, followed by South Africa, China, Indonesia, and Cambodia. Epidemiological impact projection took precedence, as 16 studies estimated the effects of vaccines on health outcomes. Additionally, 11 studies explored the costing, cost-effectiveness, and/or economic implications of vaccine implementation, emphasizing the comprehensive evaluation of benefits. The studies examined various vaccine candidates and profiles, with generic vaccines being the most common (14 studies), followed by M72/AS01E (PoD) (7 studies) and BCG revaccination (PoI) (3 studies). The primary endpoints of the generic vaccine profiles modelled with were PoD (13 studies) and PoI (5 studies) (see table 6 & 7).

**Table 4.** Study outcomes from studies modelling health impacts.

| Author, year | Country | TB vaccine | Endpoint (VE<sup>3</sup>) | Coverage | Timespan | Target population | Implementation | Projected impact |
| --- | --- | --- | --- | --- | --- | --- | --- | --- |
| Knight, 2014<sup>17</sup> | 91 LMICs | Generic vaccine | PoD (60%) | Routine vaccination: Coverage based on school attendance level.<br>Mass vaccination: 75% | 2024-2050 | Adults and adolescents | Routine vaccination: 10-year-olds<br>Mass program: 11+year olds | 17 (95% range: 11–24) million TB cases could be averted |
| Clark, 2023<sup>15</sup> | 105 LMICs | Generic vaccine | PoD (50%) | Routine vaccination: 80%<br>Mass vaccination: 70% | 2028-2050 | Adults and adolescents | Routine vaccination: 9years olds<br>Mass program: 10+ year olds. | **Routine vaccination only:**<br>8.8 million TB cases could be averted.<br>1.1 million TB deaths could be averted.<br>**Routine + one time campaign**<br>44 million TB cases could be averted.<br>5 million TB deaths could be averted.<br>**Quick scale up routine + one time campaign**<br>65.5 million TB cases averted.<br>7.9 TB deaths averted |
| Arinam-inpathy, 2022<sup>13</sup> | 24 high burden countries | Generic vaccine | PoD (60%) | South Asia: 72%<br>CAR/EU: 72%<br>Sub-Saharan Africa: 40% | 2022-2030 | Adults and adolescents | Mass vaccination: scaled up linearly | **Mass vaccination + combination other interventions<sup>4</sup>**<br>**South Asia**<br>Annual incidence rate from 325 TB cases per 100k to <75 TB cases per 100k<br>Annual mortality rate from 32 TB deaths per 100k to 7 TB deaths per 100k<br>**Sub-Saharan Africa**<br>Annual incidence rate from 205 TB cases per 100k to 75 TB cases per 100k<br>Annual mortality rate from 47 TB deaths per 100k to 8 TB deaths per 100k<br>**CAR/EU**<br>Annual incidence rate from 50 TB cases per 100k to 19 TB cases per 100k |

<sup>3</sup> Vaccine efficacy
<sup>4</sup> Improved diagnostics, routine TB services, upstream case-finding, preventive therapy, mass vaccination

|  |  |  |  |  |  |  |  |  |
| --- | --- | --- | --- | --- | --- | --- | --- | --- |
|  |  |  |  |  |  |  |  | Annual mortality rate from 5 TB deaths per 100k to <2 TB deaths per 100k |
| Fu, 2021<sup>14</sup> | 30 high RR-TB burden countries | M72/AS01E | PoD (50%) | Routine vaccination: Coverage based on school attendance level.<br>Mass vaccination: 72–76% | 2020-2035 | Adults and adolescents | Routine vaccination: 15-year-olds<br>Mass vaccination: 15+ year olds | **Vaccination only**<br>620,000 (95% credible intervals [CrI]: 516,000–867,000) RR-TB cases could be averted<br>**Vaccination + improved management of RR-TB**<br>831,000 (95% CrI: 643,000–1,170,000) RR-TB cases could be averted |
| Harris, 2020<sup>20</sup> | China, India, South Africa | BCG revaccination & M72/AS01E | PoD & Pol (70%) | Routine vaccination: 80%<br>Mass vaccination: 70% | 2025-2050 | Adults and adolescents | Routine vaccination: 9-year-olds<br>Mass vaccination: 10+ year olds | **Vaccine with POI + POD**<br>**China**<br>11.6 million (10.2 to 12.6 million) TB cases could be averted.<br>0.3 million (0.1 to 0.5 million) TB deaths could be averted.<br>**South Africa**<br>4.3 million (2.5 to 7.0 million) TB cases could be averted.<br>0.9 million (0.5 to 1.6 million) TB deaths could be averted.<br>**India**<br>4.3 million (2.5 to 7.0 million) TB cases could be averted.<br>0.9 million (0.5 to 1.6 million) TB deaths could be averted. |
| Silva, 2021<sup>23</sup> | India, Indonesia | M72/AS01E | PoD (49.7%) | Mass vaccination: 90% | 2020-2050 | Adults and adolescents | Mass vaccination: coverage reached in 10 years | **India**<br>2.3 million TB deaths could be averted.<br>**Indonesia**<br>695,000 TB deaths could be averted. |
| Weerasuriya, 2021<sup>24</sup> | China, India | Generic vaccine | PoD & Pol (50%) | Routine vaccination: 80%<br>Mass vaccination: 70% | 2027-2050 | Adults and adolescents | Routine vaccination: 9-year-olds<br>Mass vaccination: 10+ year olds | **Pre-infection and post-infection vaccine**<br>**India**<br>2.0 (UI: 1.4–4.1) million RR/MDR-TB cases could be averted<br>0.4 (0.3–0.7) million RR/MDR-TB deaths could be averted<br>**China** |
|  |  |  |  |  |  |  |  | 2.1 (UI: 1.1–2.7) million RR/MDR-TB cases could be averted<br>0.1 (0.0–0.2) million RR/MDR-TB deaths could be averted<br>**Post-infection vaccine**<br>**India**<br>1.3 (0.9–2.6) million RR/MDR-TB cases could be averted<br>0.3 (0.2–0.4) million RR/MDR-TB deaths could be averted<br>**China**<br>0.7 (0.5–0.9) million RR/MDR-TB cases could be averted<br>0.04 (0.02–0.06) million RR/MDR-TB deaths could be averted |
| Dye, 2013<sup>25</sup> | South Africa | BCG revaccination | Pol (80%) | 100% | Not provided | Adolescents | Vaccination: 15-year-olds | 17% of cases could be averted |
| Shrestha, 2017<sup>21</sup> | South Africa | Generic vaccine | PoD (60%) | Not provided | 20 years | Miners (18–60-year-olds) and (mining) labor sending community | Routine vaccination of new and repurposed miners at recruitment<br>Integration with medical examination | **Targeting miners**<br>8,090 (95% range, 3,750–13,300) TB cases could be averted<br>**Targeting (mining) labor sending community**<br>5,510 cases (95% range, 2,360–10,000) TB cases could be averted |
| Jayawardana, 2022<sup>19</sup> | South Africa | M72/AS01E | PoD (50%) | Routine vaccination: 40%<br>Mass vaccination: 60% | 2025-2050 | Adults and adolescents, PLHIV | Routine vaccination: 18–50-year-olds<br>Mass vaccination: 18–50-year-olds | **All 18–50-year-olds**<br>**Mass + routine vaccination**<br>315,256 TB cases could be averted.<br>61,718 TB deaths could be averted.<br>**2 mass vaccination campaigns**<br>490,008 TB cases could be averted.<br>96,417 TB deaths could be averted.<br>**All PLHIV adults**<br>**Mass+ routine vaccination**<br>209,524 TB cases could be averted.<br>42,143 TB deaths could be averted.<br>**2x mass vaccination**<br>367,862 TB cases could be averted.<br>73,191 TB deaths could be averted |
| Harris, 2019<sup>18</sup> | China | Generic vaccine | PoD (60%) | 70% | 2025-2050 | Adolescents and elderly | • Routine vaccination: 15-year-olds<br>• Mass vaccination: 16–19 years olds<br>• Mass vaccination: 60 64-year-olds | **Pre infection**<br>**Adolescents**<br>248,000 (214,000–292,000) TB cases could be averted<br>3,000 (2,000–5,000) TB deaths could be averted<br>**Elderly**<br>370,000 (287,000–504,000) TB cases could be averted<br>9 (5–21) TB deaths could be averted<br>**Post infection latency only**<br>**Adolescents**<br>8,000 (6,000–11,000) TB cases could be averted<br>90 (50–200) TB deaths could be averted<br>**Elderly**<br>658,000 (131,000–1081,000) TB cases could be averted<br>16,000 (2,000–45,000) TB deaths could be averted<br>**Post infection latency or recovered.**<br>**Adolescents**<br>12,000 (9,000–16,000) TB cases could be averted<br>140 (70–290) TB deaths could be averted<br>**Elderly**<br>1,295,000 (1,037,000–1,469,000) TB cases could be averted<br>33,000 (16,000–65,000) TB deaths could be averted<br>**Pre infection and post infection**<br>**Adolescents**<br>259,000 (224,000–304,000) TB cases could be averted<br>3,000 (2,000–5,000) TB deaths could be averted<br>**Elderly**<br>1,643,000 (1,403,000–1,893,000) TB cases could be averted<br>42,000 (22,000–81,000) TB deaths could be averted |
| Liu, 2017<sup>26</sup> | China | Generic vaccine | Pol (75%) | 100% | 2018-2035 | General population | Pulse vaccination of all ages (except neonatal which has its own campaign) | **Mixed vaccination**<br>The proportion of infectious population reduces to <0.002 |
| Awad. 2020<sup>27</sup> | India | Generic vaccine | PoD & Pol (50%) | 50% | 2020-2050 | People with diabetes mellitus | Not specified | **Conservative post-exposure vaccine**<br>1.7 million cases could be averted.<br>**Combination of pre- and post-exposure vaccine**<br>7.1 million TB cases could be averted |
| Shrestha, 2016<sup>22</sup> | India | Generic vaccine | PoD (60%) | Routine vaccination: 80%<br>Mass vaccination in hotspot<sup>5</sup>: 80%<br>Mass vaccination general population 14-18% | 10 years | Adults and adolescents | Routine vaccination: 10-year-olds<br>Mass vaccination of 20+year olds in hotspot or general population | **Spatially targeted vaccination (STV) program**<br>TB incidence reduction of 28%<br>**Universal targeted vaccination program**<br>TB incidence reduction of 24% |
| Clark, 2023<sup>28</sup> | India | M72/AS01E, BCG revaccination, generic vaccine | PoD & Pol (50%) | Routine vaccination: 80%<br>Mass vaccination: 70% | 2025-2050 | Adults and adolescents | Routine vaccination: 15-year-olds<br>Mass vaccination: 16+ year olds | **M72/AS01E**<br>12.7 (95% uncertainty interval: 11.0–14.6) million TB cases could be averted<br>2.0 (1.8–2.4) million TB deaths could be averted<br>**BCG revaccination**<br>9.0 (7.8–10.4) million TB cases could be averted<br>1.5 (1.3–1.8) million TB deaths could be averted |
| Renardy, 2019<sup>29</sup> | Cambodia | Generic vaccine | PoD & Pol (100%) | 80-99% | 2021-2075 | Adults and adolescents | Mass vaccination of varying age groups | Yearly, TB cases could go from 300 per 100,000 to <50 per 100,000 depending on age groups. |

\*\* LMICs: Low-income countries (LIC), and lower middle-income countries (LMIC). LICs are countries with a GNI per capita of \$1,135 or less and LMICs with a GNI per capita between \$1,136 and \$4,465. GNI per capita may differ per year. (World Bank Country and Lending Groups – World Bank Data Help Desk).
\*\*\* High burden countries: Afghanistan, Bangladesh, Burma, Cambodia, India, Indonesia, Pakistan, Philippines, Vietnam, DR Congo, Ethiopia, Kenya, Malawi, Mozambique, Nigeria, South Africa, Tanzania, Uganda, Zambia, Zimbabwe, Kyrgyz Republic, Tajikistan, Ukraine, Uzbekistan
\*\*\*\* High RR-TB high burden countries: India, Pakistan, Indonesia, Philippines, Myanmar, Bangladesh, Ethiopia, Russia Federation, Ukraine, south Africa, Mozambique, DR Congo, Zimbabwe, Nigeria, Thailand, Angola, Kenya, China, Vietnam, DPR Korea, Kazakhstan, Uzbekistan, Somalia, Peru, Kyrgyzstan, Papua New and repurposed Guinea, Tajikistan, Belarus, Republic of Moldova, Azerbaijan.
<sup>5</sup> Area with a high TB burden

**Table 5.** Main outcomes from studies modelling costing, cost-effectiveness, and economic impact.

| Author, year | Country | TB vaccine | Endpoint (VE<sup>6</sup>) | Coverage | Time span | Target population | Implementation | Projected impact |
| --- | --- | --- | --- | --- | --- | --- | --- | --- |
| Portnoy, 2023<sup>30</sup> | 105 LMICs | Generic vaccine | PoD (50%) | Routine vaccination: 80%<br>Mass vaccination: 70% | 2028-2050 | Adults and adolescents | Routine vaccination: 9-year-olds<br>Mass vaccination: 10+ year olds | **Incremental net monetary benefit for countries in which vaccination was cost effective.**<br>\$372 billion (range: \$283 to 474 billion) |
| Portnoy, 2022<sup>31</sup> | 105 LMICs | Generic vaccine | PoD (50%) | Routine vaccination: 80%<br>Mass vaccination: 70% | 2028-2080 | Adults and adolescents | Routine vaccination: 9-year-olds<br>Mass vaccination :9+ year olds | **Total absolute GDP gains in LMICs**<br>\$1618 billion (range: \$764 to 2988 billion) |
| Knight, 2014<sup>17</sup> | 96 LMICs | Generic vaccine | PoD (80%) | Routine: coverage based on school attendance level<br>Mass vaccination: 75% | 2024-2050 | Adults and adolescents | Routine vaccination: 10-year-olds<br>Mass vaccination: 11+ year olds | **Cumulated reduction (MDR)-TB treatment costs**<br>**LIC**<br>\$5.3 million<br>**LMIC**<br>\$35.6 billion<br>**UMIC**<br>\$133.4 billion |
| Portnoy, 2022<sup>16</sup> | 105 LMICs | Generic vaccine | PoD (50%) | Routine vaccination: 80%<br>Mass vaccination: 70% | 2028-2050 | Adults and adolescents | Routine vaccination: 9 years olds<br>Mass vaccination: 10+ year olds. | **Total averted costs borne by TB-affected households.**<br>\$38.9 (\$36.6–41.5) billion could be averted<br>**Averted number of households with catastrophic costs**<br>22.9 (21.4–24.5) million households facing catastrophic costs could be averted |
| Weerasuriya, 2021<sup>24</sup> | China, India | Generic vaccine | PoD & PoI (50%) | Routine vaccination: 80%<br>Mass vaccination: 70% | 2027-2050 | Adults and adolescents | Routine vaccination: 9-year-olds<br>Mass vaccination: 10+ year olds | **Total vaccination program costs**<br>**China**<br>\$41.5 (39.8–42.6) million<br>**India**<br>\$38.6 (37.1–39.9) million<br>**TB program savings**<br>**China**<br>\$5.2 (3.9–6.8) million<br>**India**<br>\$19.4 (13.0–27.2) million |

<sup>6</sup> Vaccine efficacy

|  |  |  |  |  |  |  |  |  |
| --- | --- | --- | --- | --- | --- | --- | --- | --- |
| Silva, 2021<sup>23</sup> | India, Indonesia | M72/AS01E | PoD (49.7%) | 90% | 2020-2050 | Adults and adolescents | Mass vaccination: coverage reached in 10 years | **Vaccine introduced in 2025.**<br>**India**<br>\$2 trillion losses in income could be averted.<br>**Indonesia**<br>\$680 billion losses in income could be averted |
| Weerasuriya, 2021<sup>33</sup> | China, India | M72/AS01E | PoD (50%) | 70% | 2027-2050 | Adults and adolescents | Mass vaccination: 10+ year olds<br>Mass vaccination: various age groups | **Country-specific willingness to pay threshold.**<br>**India**<br>\$21 billion is the willingness to pay for a vaccine program.<br>\$5 billion for vaccinating the highest efficiency group (50-59y)<br>**China**<br>\$15 billion is the willingness to pay for a vaccine program.<br>\$6 billion for vaccinating the highest efficiency group (60-60y) |
| Harris, 2022<sup>32</sup> | India, South Africa | M72/AS01E | PoD (50%) | Routine vaccination (10years old): 80%<br>Routine vaccination (15years old): 80%<br>Routine vaccination (18 years old): 50% | 2025-2050 | Adolescents | Routine vaccination: 10-year-olds<br>Routine vaccination: 15-year-olds<br>Routine vaccination: 18-year-olds | **Pre- and post-infection efficacy; 18-year-olds, 50% coverage**<br>**India**<br>\$-135.1 (-324.4, 19.6) cost per DALY averted from societal perspective.<br>**South Africa**<br>\$290.1 (58.3, 723.1) cost per DALY averted from societal perspective |
| Clark, 2023<sup>28</sup> | India | M72/AS01E, BCG revaccination, Generic vaccine | PoD & PoI (50%) | Routine vaccination: 80%<br>Mass vaccination: 70% | 2025-2050 | Adults and adolescents | Routine vaccination: 15-year-olds<br>Mass vaccination: 16+ year olds | **Annual incremental program cost**<br>**BCG revaccination**<br>US\$23 million<br>**M72/AS01E**<br>\$190 million<br>**ICER (US\$/DALY averted) from a societal perspective.**<br>**M72/AS01E**<br>\$139 (77–229) per DALY averted<br>**BCG revaccination**<br>\$17 (cost-saving<71) per DALY averted |
| Dye, 2013<sup>25</sup> | South Africa | BCG revaccination | PoI (80%) | 100% | - | Adolescents | Vaccination of 15-year-olds | \$52 –\$4540 per DALY averted |
| Jayawar-dana,<br>2022<sup>19</sup> | South<br>Africa | M72/AS0<br>1E | PoD (50%) | • Routine vaccination: 40%<br>• Mass vaccination: 60% | 2025-2050 | Adults and adolescents, PLHIV | • Routine vaccination: 18–50-year-olds<br>• Mass vaccination: 18–50-year-olds | **Most cost-effective strategy: 2 mass vaccination campaigns 18–50-year-olds**<br>\$184 million reduced TB related costs<br>**Total incremental cost**<br>\$417 (IQR 400–433 million) million |

\*\* LMICs: Low-income countries (LIC), and lower and upper middle-income countries (LMIC). LICs are countries with a GNI per capita of \$1,135 or less and LMICs with a GNI per capita between \$1,136 and \$4,465. Upper middle-income economies are those with a GNI per capita between \$4,466 and \$13,845. GNI per capita may differ per year (World Bank Country and Lending Groups – World Bank Data Help Desk).

**Table 6.** Study characteristics and main measurement outcomes from epidemiological impact studies by country.

|  | LMICs | India | South Africa | China | Indonesia | Cambodia |
| --- | --- | --- | --- | --- | --- | --- |
| **Nr. studies** | 4 | 6 | 4 | 4 | 1 | 1 |
| **Vaccine type** |  |  |  |  |  |  |
| **BCG** | 0 | 2 | 2 | 1 | 0 | 0 |
| **M72/AS01E** | 1 | 3 | 2 | 1 | 1 | 0 |
| **Generic vaccine** | 3 | 4 | 1 | 3 | 0 | 1 |
| **Endpoint generic vaccines** |  |  |  |  |  |  |
| **POI** | 0 | 3 |  | 2 | 0 | 1 |
| **POD** | 3 | 3 | 1 | 2 | 0 | 1 |
| **POR** | 0 | 0 | 0 | 0 | 0 | 0 |
| **Target population** |  |  |  |  |  |  |
| **Adults and adolescents** | 4 | 5 | 2 | 2 | 1 | 1 |
| **Other** | 0 | 1<sup>9</sup> | 3<sup>10</sup> | 2<sup>11</sup> | 0 | 0 |
| **Measurement of outcomes** |  |  |  |  |  |  |
| **Incidence** | 5<sup>12</sup> | 5<sup>9</sup> | 4 | 4<sup>9</sup> | 0 | 1 |
| **Mortality** | 3 | 4<sup>9</sup> | 2 | 3<sup>9</sup> | 1 | 0 |

**Table 7.** Study characteristics and main measurement outcomes from studies modelling costing, cost-effectiveness, economic impacts by country.

|  | LMIC's | India | South Africa | China | Indonesia |
| --- | --- | --- | --- | --- | --- |

<sup>9</sup> People with diabetes mellitus
<sup>10</sup> Miners and labour sending community, PLHIV, adolescents only.
<sup>11</sup> General population, adolescents +elderly
<sup>12</sup> Of which 1 focuses on RR-TB

|  |  |  |  |  |  |
| --- | --- | --- | --- | --- | --- |
| **Nr. Studies** | 4 | 5 | 3 | 2 | 1 |
| **Vaccine type** |  |  |  |  |  |
| **BCG** | 0 | 1 | 1 | 0 | 0 |
| **M72/AS01E** | 0 | 4 | 2 | 1 | 1 |
| **Generic vaccine** | 4 | 2 | 0 | 1 | 0 |
| **Endpoint generic vaccines** |  |  |  |  |  |
| **POI** | 0 | 2 | 0 | 1 | 0 |
| **POD** | 4 | 5=1 | 0 | 1 | 0 |
| **POR** | 0 | 0 | 0 | 0 | 0 |
| **Target population** |  |  |  |  |  |
| **Adults and adolescents** | 4 | 4 | 1 | 2 | 1 |
| **Other** | 0 | 1<sup>13</sup> | 2<sup>14</sup> | 0 | 0 |
| **Measurement outcomes** |  |  |  |  |  |
| **GDP gains** | 1 | 0 | 0 | 0 | 0 |
| **Cost-effectiveness** | 2 | 2 | 2 | 1 | 0 |
| **TB vaccination program cost** | 0 | 3 | 0 | 1 | 0 |
| **Threshold willingness to pay** | 0 | 1 | 0 | 1 | 0 |
| **Averted cost per income quintile** | 1 | 0 | 0 | 0 | 0 |
| **Full-income gains** | 0 | 1 | 0 | 0 | 1 |

### Epidemiological impact

Epidemiological impact studies were mostly conducted in India (6) or in LMICs/various high burden countries in general (4), China (4), and South Africa (4). Vaccine candidates and profiles modelled with were generic vaccines (11), M72/AS01E (5), and BCG revaccination (3). Of the studies modelling with a generic vaccine, 9 studies used a PoD endpoint, 5 used PoI, and none used PoR. Most studies (11) focused on the general adult/adolescent population, 1 on adolescents only, 1 on the general population, 1 on miners, 1 on people with diabetes mellitus (DM), and one within a geographic hotspot with a high TB incidence (see table 6).

Overall, the epidemiological impact of a new and repurposed PoD TB vaccine was found to be substantial compared to a no TB vaccine scenario (beyond BCG vaccination in neonates), with the highest health gains projected within the groups/countries with the highest TB burden (see Table)^13–17^. Based on studies focusing on a selection of LMICs, the delivery strategy with the highest projected epidemiological impact consisted of routine vaccination with an instantly rolled out mass vaccination campaign, and concurrent with other public health interventions such as improved diagnostics, treatment, and reduced delay in health-seeking^13–17^. A new or repurposed PoD TB vaccine with an efficacy of 60% was found to help reach the End TB targets if a minimal coverage of 72% could be achieved in South Asia, Central Asia Republics and Europe (CAR/EU) and 40% in Sub-Saharan countries. This difference for Sub-Saharan countries is due to the projected impact of TPT uptake in PLHIV^13^.

Country-specific studies revealed differences in impact as vaccine characteristics, target population and implementation strategy, timing of roll out, TB burden, and vaccination coverage rates vary. In China, a PoD vaccine targeting older people, protective in both mycobacterium tuberculosis (Mtb.) infected and recovered populations, could deliver the highest impact^18^. In South Africa, Jayawardena et al.^19^ projected the largest epidemiological impact with a strategy implementing a sequence of two mass vaccination campaigns and a second South African study found that a PoD vaccine could be most impactful if effective in both uninfected and infected people, and safe for use among PLHIV^20^. Moreover, Shrestha et al.^21^ found a greater impact of a miner-targeted vaccine strategy compared to a community-targeted strategy in associated labour sending communities in South Africa. This was due to the higher concentration of adult men in the miner target population. In India, Silva et al. found that a vaccination program’s effectiveness targeting high incidence geographic areas depends on interaction of populations between geographic areas. Vaccination of people in a well-connected area with high TB incidence could reduce incidence 1.6-fold or more^22^. Another study showed in Indonesia and India that vaccinating either all ages or 18-49 year olds could bring substantial health and economic gains, and both health and economic gains would be more substantial with vaccinating all age groups^23^.

### Costing, Cost-effectiveness, and/or Economic impact

Eleven studies investigated costing, cost-effectiveness, and economic impacts (see table 5 & 7). The distribution of studies included general LMICs (4) and studies in India (5), South Africa (3), China (2), and Indonesia (1). Across the studied countries, the M72/AS01E vaccine candidate was most frequently studied in India (4 studies), whereas generic PoD vaccines were investigated across India (5), South Africa (3), China (2), and Indonesia (1). Costing, cost-effectiveness, and economic measurement outcomes differed across studies, with cost-effectiveness estimated for all except one country (Indonesia). Overall, adults and adolescents from the general population were the most frequently modelled target populations.

In studies focused on LMICs, adult/adolescent TB vaccine strategies could be economically beneficial compared to situations without new and repurposed TB vaccines. However, this is dependent on the TB burden, prevalence of HIV co-infection, vaccine program approaches, and ultimately the efficacy and duration of protection of the vaccine^16,17,30,31^. An adult/adolescent TB vaccine was found to be cost-effective in most countries and cost-saving in 58 LMICs^30^, with higher TB incidence countries gaining the most benefit. Furthermore, an adult/adolescent vaccine program could reduce catastrophic costs by 40%, with the highest gains in the lowest income quintiles^16^.

Country-specific estimates also illustrate that costing, cost-effectiveness, and economic impact is dependent on vaccine characteristics, TB burden, country-specific context and thresholds for cost-effectiveness, coverage, and timing of a program. In India, M72/AS01E is 7-times more cost-effective than BCG revaccination despite cost-effectiveness being evident across all scenarios^28^. Additionally, Harris et al. found that only a vaccine which includes efficacy in uninfected people was cost-effective. This was due to the epidemic in India and their specific thresholds for cost-effectiveness^32^. In South Africa, a PoD vaccine could be highly cost-effective and even the least effective PoD vaccine scenario (80% coverage of 10 year olds, post-infection protection) remained below the cost-effectiveness threshold^32^. The inclusion of HIV-related costs was a relevant contextual factor in South Africa as mortality caused by TB among PLHIV will be prevented by the vaccine. In China, Harris et al. found that a PoD vaccine effective in both TB infected and uninfected people could be most cost-effective based on RR-TB reductions^24^. Another study by Harris et al. found that in India and South Africa, the costs of vaccinating 18 year olds with 50% coverage was equal to the costs of vaccinating 15 year olds with 80% coverage^32^. Lastly, Silva et al. demonstrated that for India and Indonesia, a delay in implementation from 2020 to 2025 and the subsequent loss in economic impact could be recovered by vaccinating all ages instead of only a specific age group (18-49 year olds)^23^.

### Acceptability

One study assessed the acceptability of TB vaccine implementation in India, China, and South Africa^34^. This article evaluated acceptability among high level vaccine policy and TB experts (Table 8. Factors influencing acceptability and demand of a new and repurposed TB vaccine). In all three countries, barriers included TB-related stigma affecting coverage during a mass vaccination campaign as well as affordability. Barriers to uptake specific to China included vaccine hesitancy and perceived level of protection. In India, experts perceived negative publicity of side effects as barrier to acceptability by the public and were hesitant about BCG revaccination resulting from previous trials showing no effects among Indian children. South African specific barriers included vaccine hesitancy and the need for a second dose of a vaccine candidate. The facilitators identified across countries included low price per dose, and public awareness of TB disease and the new and repurposed TB vaccine. Moreover, routine vaccination strategies, engaging communities/civil societies, and familiarity with the vaccine (in the case of BCG revaccination) were considered facilitators in the Chinese and South African contexts.

**Table 8.** Factors influencing acceptability and demand of a new and repurposed TB vaccine^34^.

|  | India | China | South Africa |
| --- | --- | --- | --- |
| **Beliefs and attitudes** | - | - | - |
| **Barriers** | • Stigma<br>• Negative publicity of side effects<br>• High cost<br>• Implementing the BCG revaccination | • Stigma<br>• Vaccine hesitancy<br>• High cost<br>• Low perceived level of protection | • Stigma<br>• Vaccine hesitancy<br>• The need for a second dose<br>• High cost |
| **Enablers /Facilitators** | • High awareness, especially among elderly<br>• Low price | • Routine vaccination<br>• Vaccination campaign Integrated within national immunization program.<br>• High awareness/health education<br>• Community engagement<br>• Low price<br>• Familiarity with the BCG vaccine if BCG revaccination is implemented | • Routine vaccination<br>• Strong political commitment<br>• Lessons learned from the COVID-19 vaccination program.<br>• Civil society engagement<br>• Low price<br>• Familiarity with the BCG vaccine if BCG revaccination is implemented |

### Implementation feasibility

Five articles contained information on key considerations for implementation feasibility, including implementation strategies of an adult/adolescent TB vaccine in LMIC, and health system readiness ^19,21,22,34,35^. shows the number of implementation feasibility studies by country. Three countries were investigated, with South Africa being the most represented.

### Implementation strategies by target population

Five articles contained information related to country-specific implementation strategies, in the context of China, India, and South Africa (Table 9)^19,21,22,34,35^. Various strategies for TB vaccine implementation were identified, which are summarized in table 10. Target populations that were similar across the countries were adolescents, adults, socially vulnerable groups, PLHIV and other people with immunocompromised systems, and healthcare workers^34,35^. The elderly and high risk contacts were target populations evaluated in India and China^34^ and in South Africa the mining-community was evaluated as a target population^21^. In the Indian context, a focus on high-risk populations by targeting high incidence geographic ‘hotspots’ was also assessed^22^.

**Table 9.** Characteristics of studies evaluating facets related to implementation feasibility by country.

|  | India | South Africa | China |
| --- | --- | --- | --- |
| **Nr studies feasibility** | 2 | 4 | 1 |
| **Vaccine type** |  |  |  |
| BCG | 1 | 2 | 1 |
| M72/AS01E | 1 | 2 | 1 |
| Generic vaccine | 1 | 2 | 0 |
| **Endpoint generic vaccines** |  |  |  |
| POI | 0 | 1 | 0 |
| POD | 1 | 2 | 0 |
| POR | 0 | 0 | 0 |
| **Target population** |  |  |  |
| Adolescents | 1 | 1 | 1 |
| Adults | 1 | 1 | 1 |
| Elderly | 1 | 0 | 1 |
| Socially vulnerable groups | 1 | 1 | 1 |
| PLHIV | 1 | 1 | 1 |
| Immunocompromised groups | 1 | 1 | 1 |
| Healthcare worker | 1 | 2 | 1 |

<sup>13</sup> Adolescents only
<sup>14</sup> Adolescents only, PLHIV adults

|  |  |  |  |
| --- | --- | --- | --- |
| Other | 2<sup>15</sup> | 1<sup>16</sup> | 1<sup>17</sup> |
| Implementation strategies | 2 | 4 | 1 |
| Health system readiness | 1 | 1 | 1 |

**Table 10.** Overview of studies assessing implementation strategies.

| Author, year | Country | TB vaccine | Endpoint | Time span | Target population | Implementation strategies |
| --- | --- | --- | --- | --- | --- | --- |
| Pelzer, 2022<sup>34</sup> | China, India, South Africa | M72/ BCG revaccination | Pol/PoD | 2025-2030 | Adolescents<br>Adults<br>Elderly<br>Socially vulnerable groups<br>PLHIV<br>Immunocompromised<br>HCW<br>Other | Routine vaccinations<br>Targeted vaccination programs<br>Mass campaigns<br>Integration into existing healthcare systems |
| Hatherill, 2016<sup>35</sup> | South Africa | Generic vaccine & BCG revaccination | Pol/PoD | Not specific | Healthcare workers | Integration within the occupational health infrastructure |
| Shrestha, 2016<sup>22</sup> | India | Generic vaccine | PoD | Not specific | Population living in geographic high incidence TB ‘hotspots’ | Spatially targeted vaccine program |
| Shrestha, 2017<sup>21</sup> | South Africa | Generic vaccine | PoD | Not specific; 20 years | Mining population | Routine vaccination of new and repurposed miners at recruitment<br>Integration with medical examination |
| Jayawardana, 2022<sup>19</sup> | South Africa | M72 | PoD | 2025-2050 | Adults/adolescents and PLHIV adults | Mass vaccination combined with routine annual vaccination of 18-year-olds, two mass campaigns |

### Health system readiness

Two articles discussed health system readiness for new and repurposed TB vaccine implementation in India, China, and South Africa (Table 11. Overview of studies assessing health system readiness)^34,35^. For all three countries, there was a described need for an extensive monitoring and surveillance system to track potential adverse events. In India, it was mentioned that a mass vaccination campaign could be hindered by new and repurposed cold chain requirements, while routine vaccination could be more easily integrated within the existing public health system. Additionally, testing for Mtb. infection was considered too expensive and unfeasible in India. In South Africa, testing for Mtb. infection and the high prevalence of HIV infection-which may limit which vaccines could be safely used-were also potential challenges. In China, storage capacity was an additional factor that could delay effective implementation, yet the current procurement and supply chain could manage the integration of a new and repurposed vaccine program. In South Africa and China, insufficient staff capacity could hinder any future vaccination campaign.

**Table 11.**
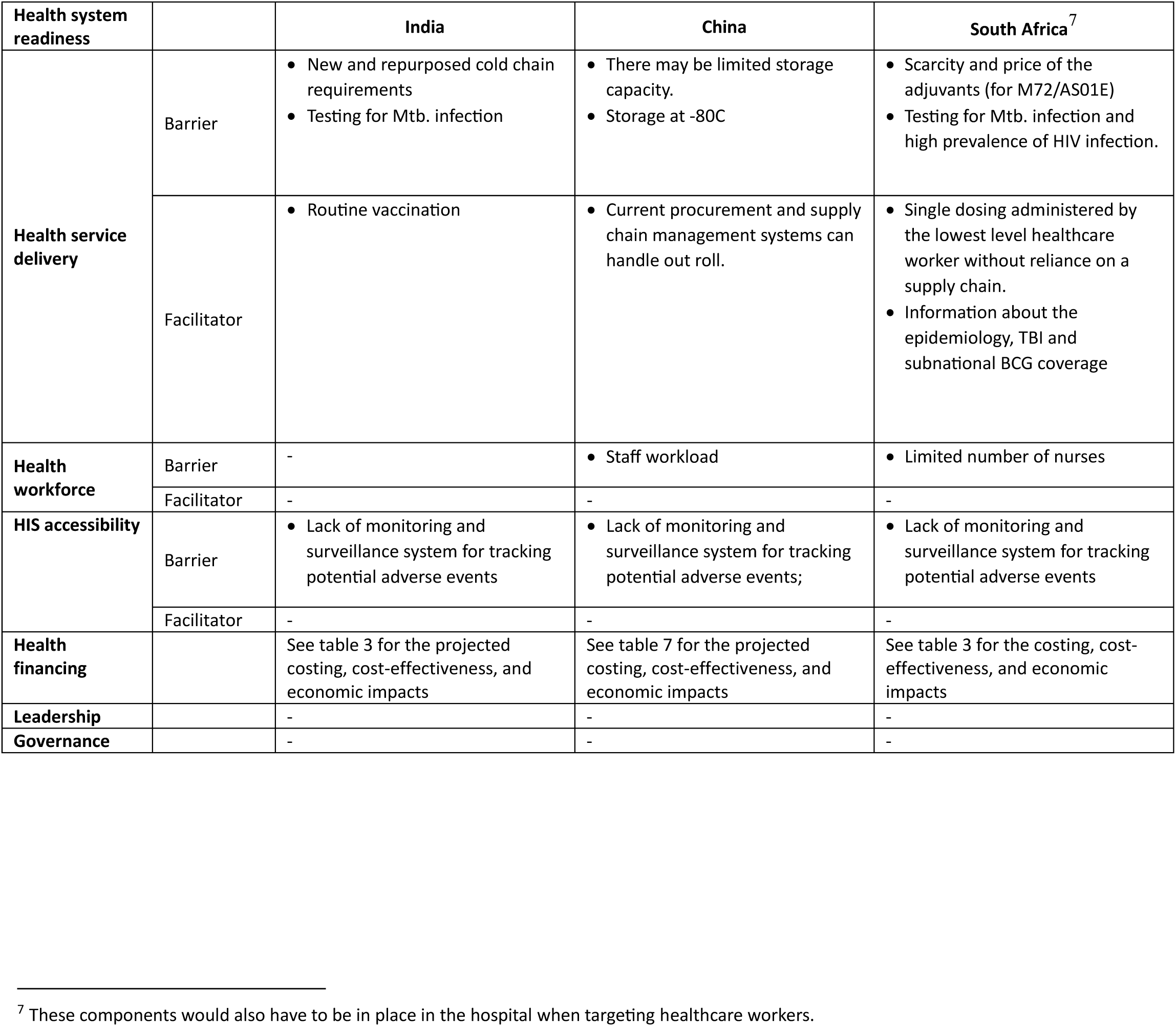
Overview of studies assessing health system readiness.

## Discussion

In this scoping review, we found that there is limited body of evidence on implementation potentials of new and repurposed TB vaccines among adults and adolescents. Most studies assessed epidemiological or costing/cost-effectiveness and/or economic impacts of new and/or repurposed TB vaccines followed by implementation feasibility while only one study contained information regarding acceptability among high level stakeholders. The Sub-Saharan African region was underrepresented in all the studies assessed in this review. While our findings clearly highlight the availability of evidence that points toward potential impacts of new and repurposed TB vaccines among adults and adolescents, it also underscores the need for broader geographic and target population representation in the research field.

While the epidemiological studies showed that new and repurposed TB vaccines may avert millions of TB cases and TB deaths^15^, the estimated impacts differed per country and were dependent on vaccine characteristics, target population and implementation strategy, the timing of roll out, the local TB burden, and vaccination coverage^18–23^. Costing, cost-effectiveness, and economic studies showed that in a majority of countries, and remarkably, in 58 LMICs, an adult/adolescent vaccine program could potentially not only be cost-effective but even yield cost-savings outcomes^30^. Vaccine characteristics, TB burden, cost-effectiveness, coverage, and timing all influence costing, cost-effectiveness, and economic-related outcomes^15–17,23,24,28,30,32^. Vaccine candidates and profiles modelled with included generic vaccines, M72/AS01E, and BCG revaccination like candidates. Vaccine efficacies ranged from 50%-100%, with coverage rates between 40%-100%. Most studies focused on PoD, some on PoI, and the target population was mainly general adults and adolescents. The study investigating factors influencing acceptability of new and repurposed TB vaccines in India, South Africa, and China showed that countries may differ in their preference for a vaccine candidate and barriers/benefits. In all three countries, common barriers included TB-related stigma affecting coverage during a mass vaccination campaign as well as affordability. Facilitators identified across countries were low price per dose, and public awareness of TB disease and the new and repurposed TB vaccine^34^. Lastly, studies focusing on implementation feasibility showed that strategies could include mass campaign(s), integrated within existing routine vaccine programs, and targeted campaigns specifically for-for example-miners, elderly, PLHIV, and people with immunocompromising conditions. This will be dependent on country specific context in terms of burden and health system readiness and capabilities^19,21,22,34,35^.

While this scoping review identified limited information on implementation feasibility, prior research confirms the few findings and underscores the need for more research in this field to ensure effective implementation, once a new and/or repurposed TB vaccine is available. Vaccine hesitancy, lack of availability of monitoring and surveillance systems, affordability and funding/cost of delivery, and storage requirements were also challenges identified in previous studies^36–38^. Additionally, estimating target population size, slow roll-out of the vaccination campaign, accessibility, and wars/conflicts were additional challenges mentioned in reviews specifically focusing on LMICs^38,39^. A holistic and timely implementation planning process, expanding access, reducing out-of-pocket costs, one-dose vaccine programs, extensive community involvement, and implementation of programs in schools could circumvent some of these challenges^36,38,39^. Notably, as assessed in the review by Wang et al., introducing a new vaccine may also have broader positive effects on the general health system^40^. They found that besides reductions in the disease burden, improvements in disease and adverse events surveillance, training, cold chain and logistics capacity and injection safety were documented as beneficial effects. However, they also emphasize that opportunities for strengthening the broader health system were consistently missed during out roll^40^.

Research in LMICs other than India, China, and South Africa was lacking, which is especially important due to three aspects. Firstly, most TB cases and deaths are concentrated in LMICs with eight countries comprising of around two-thirds of the burden worldwide^41^. Secondly, epidemiological variations among countries influence TB vaccine implementation. Therefore, factors such as TB/HIV prevalence, tuberculosis infections (TBI) rates, BCG vaccine coverage, and multi-drug resistant (MDR)-TB burden must be considered. Thirdly, while BCG revaccination and M72/AS01E vaccine candidates are advanced, they may not be suitable for all contexts and populations.

Further clinical research is needed to identify which vaccines are likely effective and approvable. This will guide the characteristics to consider in future modelling. Currently, research primarily relies on expert opinions, but it lacks essential acceptability assessments among end users. Although cross-country representation is significant, we need to consider necessary specific target populations within each country. As shown in this review, different countries have varying target populations, each with distinct estimated impacts and coverage levels. It is vital to expand research to include other LMICs, various target populations, and late-stage vaccine candidates. This broader approach will enhance our understanding of vaccine characteristics, leading to better decision-making and equitable distribution.

Understanding the TB burden and TB infection rates within various target populations across countries is paramount for optimizing the potential benefits of new and/or repurposed TB vaccines. This foundational knowledge serves as the basis upon which tailored vaccine implementation strategies can be formulated. Crucially, the development of implementation strategies should be guided by feasibility and equity considerations. These strategies should be linked to the TB burden in specific regions, considering contextual factors and health disparities that impact the effective deployment of TB vaccines. The experiences of China, India, and South Africa, which have chosen routine vaccination as a practical implementation strategy, underscore the feasibility of integrating new and repurposed vaccines into existing vaccination programs. Consequently, we recommend future TB vaccine preparedness research to investigate country-specific TB/TBI rates within target populations.

Further examination of factors such as stigma, accessibility, availability, vaccine hesitancy, and vaccine confidence, along with an exploration of the mechanisms, facilitators, and barriers influencing these aspects, holds pivotal significance. These determinants exert a direct impact on the acceptance, adoption, and uptake of novel or repurposed TB vaccines. Considerations like community beliefs and attitudes, equitable representation, and individual choices regarding vaccination are subsequently essential for informing the landscape of acceptability and feasibility. Regrettably, the existing body of research exhibits a conspicuous gap in knowledge, as no prior studies have delved into these critical topics. Gallagher et al.^42^ and McClure et al.^43^ underscore the significance of addressing hesitancy and rumors, as these factors can detrimentally affect vaccine programs on a broader scale. The absence of research in this domain underscores the urgent necessity for comprehensive investigations aimed at better comprehending the dynamics that shape individuals’ attitudes, perceptions, and behaviors towards TB vaccines.

Furthermore, it is of utmost importance to assess the readiness of healthcare systems in (LMICs for the seamless integration of new and repurposed TB vaccines. Experience from the implementation research of COVID-19 vaccination has highlighted the critical importance of health system readiness^44^. This readiness is essential to mitigate delays and prevent losses in the implementation of TB vaccines and should encompass an evaluation of healthcare infrastructure, capacity, and potential obstacles, enabling the development of well-informed strategies that facilitate successful implementation. See table 12 for an overview of the prioritized gaps for future research to guide new and repurposed TB vaccine implementation.

**Table 12.** Prioritized gaps for future research to guide new and repurposed TB vaccine implementation.

|  | Geographical | Vaccine related | Data/ Tools | Other |
| --- | --- | --- | --- | --- |
| **Overall** | Country specific evidence | Other pipeline candidates<br>PoI, PoR vaccines |  | After 2050, i.e., long-term impact<br>Various target populations |
| **Epidemiological impact** | Global, country specific evidence | Other pipeline candidates<br>PoI vaccines,<br>Validated VE<br>Impact without prior BCG | Country validated estimates.<br>Target population estimates | Combination with other TB interventions<br>Country specific target populations |
| **Economic impact** | Global, country-specific context | PoD, PoI, PoR vaccines | thresholds for cost-effectiveness | Coverage<br>Timing of a program,<br>Combination with other TB intervention country specific target populations |
| **Implementation feasibility** | Global, country specific evidence | PoD, PoI, PoR vaccines | Implementation strategies<br>Health system readiness assessment<br>Acceptability<br>Health equity | Geographic analysis of accessibility to vaccine locations |
| Acceptability | Global, country specific evidence | PoD, PoI, PoR vaccines | Attitudes, beliefs, perceptions, preferences, decision-making metrics | Acceptability among target populations<br>Preferences of target populations, including in comparison to other TB interventions |
| Implementation strategies | Global, country specific evidence | PoD, PoI, PoR vaccines | Implementation strategies | Country specific target populations |

<sup>15</sup> High-risk contact, high resource areas and occupational groups, spatially targeted vaccines in geographic hotspots
<sup>16</sup> Mining population
<sup>17</sup> High-risk contact, high resource areas and occupational groups

|  |  |  |  |  |
| --- | --- | --- | --- | --- |
| Health system readiness | • Global, country specific | • PoD, PoI, PoR vaccines | • Validated assessment tool<br>• Country validated essential indicators | • Country specific target populations<br>• Geographic analysis of catchment areas of expected vaccine locations |

Our study has limitations. Notably, we conducted a scoping review, which is distinct from a comprehensive systematic review. While we employed diverse methods to comprehensively map the existing literature and identify gaps, it is possible that some relevant studies may have been inadvertently omitted. Nevertheless, our primary aim was to present a comprehensive overview of the current state of the field, rather than striving for an exhaustive compilation. Additionally, our scope was exclusively focused on TB vaccine implementation. Over recent years, however, much experience has been gained with implementation of vaccinations for adults and adolescents e.g., COVID-19, Hepatitis B, and HPV. These lessons learned would be very relevant and valuable to the implementation of a new or repurposed TB vaccine in the same age groups. Despite these limitations, it is the first review to comprehensively map the existing literature on the implementation and impacts of new and repurposed TB vaccines. This provides stakeholders with a clear overview of the current body of knowledge, enhancing the understanding of stakeholders regarding the current landscape and aiding them in prioritizing future research in this area.

## Conclusion

With several TB vaccine candidates in the pipeline, there is an urgent need for research on the global implementation of effective TB vaccines among adult and adolescent populations, including assessment of economic and epidemiological impacts in LMICs. Country-specific evidence on TB vaccine implementation is also imperative but is currently lacking. Addressing these gaps will be needed to prepare countries for novel TB vaccine implementation, upon approval, to ensure uptake through the design of optimal and equitable vaccine implementation strategies. Future studies should focus on generating evidence on understudied areas such as key factors influencing acceptability and implementation feasibility. Furthermore, geographic and population diversity should be seriously considered when planning further studies to understand TB vaccine implementation strategies.

## Author contributions

JSB, PTP, DJ and AG conceived the study and wrote the protocol. JSB performed a systematic search and screened and identified studies. PTP validated and verified the search, screening, and identification. JSB and PTP made final decisions regarding study inclusion. JSB and PTP wrote the first draft of this manuscript. RB, DJ, and AG contributed to data analysis and interpretation and manuscript review and revision. ADK AM, and RL reviewed the protocols and revised the manuscript. All authors have read and edited the drafted manuscript. All authors approved the final version of the manuscript.

## Acknowledgements

This review is executed under the SMART4TB project. SMART4TB is made possible by the generous support of the American people through the United States Agency for International Development (USAID) and is implemented under cooperative agreement number 7200AA20CA00005. The consortium is managed by prime recipient, Johns Hopkins University.

## Data Availability

This is a review of published (preprint) research articles. All studies reviewed are available in either of the following databases: PubMed, Medrxiv, and PLOS. The lists of search strings can be found in Annex A- search strings. Additionally, all relevant data used in the review are in the manuscript and Annex B-supplementary tables.

## Footnotes

1 LMICs includes low-, lower-middle, and upper-middle income countries depending on the Gross National Income (<u>Low-and Middle-</u> <u>income Countries - List | Wellcome</u>)

2 A separate review is assessing lessons learnt from vaccine introductions, other than new and repurposed TB vaccines.

## Annex A – Search Strings

| database | concept | Search string | filters |
| --- | --- | --- | --- |
| PubMed | #1 tuberculosis | “Tuberculosis”[MeSH Terms] |  |
|  | #2 Tuberculosis vaccination | “Tuberculosis vaccines” [MeSH Terms] OR “tb vaccin*”[Title/Abstract] OR “tuberculosis vaccin*”[Title/Abstract] |  |
|  | #3 Vaccine preparedness | “Immunization Programs”[MeSH Terms] OR “Vaccination Refusal”[MeSH Terms] OR “Vaccination Coverage”[MeSH Terms] OR “Vaccination Hesitancy”[MeSH Terms] OR “implement*”[Title/Abstract] OR “strateg*”[Title/Abstract] OR “barrier*”[Title/Abstract] OR “enabler*”[Title/Abstract] OR “factor*”[Title/Abstract] OR “health system*”[Title/Abstract] OR “prepare*”[Title/Abstract] OR “readiness”[Title/Abstract] OR “roll out”[Title/Abstract] OR “impact”[Title/Abstract] OR “effect*”[Title/Abstract] OR “intro*”[Title/Abstract] OR “uptake”[Title/Abstract] OR “accept*”[Title/Abstract] OR “availab*”[Title/Abstract] OR “access*”[Title/Abstract] OR “feasib*”[Title/Abstract] OR “human resources*”[Title/Abstract] OR “capacity*”[Title/Abstract] OR “coordinat*”[Title/Abstract] OR “training*”[Title/Abstract] OR “cost*”[Title/Abstract] OR “financ*”[Title/Abstract] OR “govern*”[Title/Abstract] OR |  |
|  |  | “regulat*”[Title/Abstract] OR “service deliver*”[Title/Abstract] OR “scale up*”[Title/Abstract] OR “scalib*”[Title/Abstract] OR “hesitan*”[Title/Abstract] OR “facilitat*” [Title/Abstract] |  |
|  | Combined with exclusion | #1 AND #2 AND #3 AND “Humans” [Mesh] NOT “Clinical Study” [Publication type] NOT “Infant” [Mesh] NOT “Child, Preschool” [Mesh] NOT “Immunity” [Mesh] | Year: 2013-2023 |
| Medrvix | Tuberculosis vaccin | for term “tuberculosis vaccin” and abstract or title “adolescent adult” (match any words) and posted between "1<sup>st</sup> of Jan 2013 and 30<sup>th</sup> of Apr, 2023" | Year: 2013-2023 |
| Plos Global public health | Vaccine and tuberculosis | ((abstract:vaccin) AND abstract:tuberculosis) | Year: 2013-2023 |

## Annex B – Supplementary tables

## Notes

### Competing Interest Statement

The authors have declared no competing interest.

### Funding Statement

We are funded by SMART4TB, the grant number is 7200AA20CA00005. The funders had no role in study design, data collection and analysis, decision to publish, or preparation of the manuscript.

### Author Declarations

KNCV Ethics Review Board reviewed and approved of the review as it meets international ethics standards and does not pose any ethical issues.

